# Co-production of a Neurodiversity-Affirmative Anxiety Intervention for Autistic Children

**DOI:** 10.1101/2023.07.05.23292219

**Authors:** Tasha Cullingham, Una Rennard, Cathy Creswell, Damian Milton, Karen Leneh Buckle, Lucie Godber, Kate Gordon, Michael Larkin, Jonathan Green

**Affiliations:** CAMHS, Manchester University Foundation Trust; Expert by Experience; Departments of Experimental Psychology and Psychiatry, University of Oxford; Tizard Centre, University of Kent; Department of Psychology, Communication and Human Neuroscience, Faculty of Biology, Medicine and Health, University of Manchester; Department of Neuropsychology, Berkshire Healthcare NHS Foundation Trust; College of health and Life Sciences, Aston University

**Keywords:** Autism, Co-production, Neurodiversity, CBT, Child anxiety, Parent-led interventions

## Abstract

**Background:** Mental health difficulties are common for autistic people; however, few interventions have been co-produced with the autistic community. Mental health interventions constructed with a ‘non-autistic lens’ likely miss key understandings from autistic experiences and priorities of the autistic community. Additionally, the style and aims of intervention may be prone to unconscious bias around the pathologisation of autism. Currently, there are limited methodological and practical examples of how to rigorously co-produce mental health interventions with autistic people. This paper details the methodology and processes of co-adapting an intervention for autistic children with anxiety problems. Providing a worked example of co-producing a neurodiversity-affirmative mental health intervention which reflects autistic, parental, academic, and clinical, experience and expertise.

**Methods:** We co-adapted content from a brief, parent-led, CBT approach for non-autistic children with anxiety problems to meet the needs of autistic children and their parents. The adaptation for autistic children was co-constructed using processes and strategies adopted from Experience-Based Co-Design (EBCD). The research team, comprising autistic and non-autistic members, worked alongside an expert reference group (ERG). The ERG comprised parents (autistic and non-autistic) of autistic children with anxiety problems, autistic adults with experience of anxiety problems, and clinicians with experience supporting autistic children with mental health difficulties. Data were obtained from qualitative research interviews with autistic children with anxiety problems and parents. These data were considered reciprocally by the research team and the ERG.

**Results:** The resulting intervention includes a neurodiversity-affirmative perspective that considers how anxieties for autistic children can emerge from being neurodivergent in a neurotypical world integrated with traditional CBT techniques and understandings of anxiety.

**Conclusion:** Successful co-production can help to integrate multiple theoretical backgrounds and result in the creation of interventions that are potentially acceptable to clinicians, autistic people and their family members.

## Why Autistic Perspectives are Required

Co-production has many different definitions but at its core is a collaborative partnership between key stakeholders often including patients, the public, clinicians, and researchers. Within true co-production the inherent power balances are dismantled and the power and responsibility are shared. Among the many benefits of co-production, it can provide a place early in the research process to work through conflicting opinions between stakeholders, and thus contribute to more acceptable and relevant research outputs. This is of particular relevance within the field of autism where the clinical concept of autism and neurodiversity paradigm are at epistemological odds, leading to a divisive split.

The clinical concept of autism has historically evolved within a framework of identifying neurodevelopmental differences purely as impairments or deficits; a formulation now challenged by the neurodiversity paradigm and many in the autistic and wider autism community. The neurodiversity movement acknowledges the inherent value of all people and the importance of the human population containing differing neurological phenotypes. Focusing on acceptance of difference and variation, rather than ‘fixing’ deficits. Leadbitter et al. (2021) argue that a neurodiversity-informed approach is essential for ethical and respectful interventions for autistic children. This is of particular importance as the deficit-focused conceptualisation of autism can be highly damaging to the well-being of autistic people (Botha & Frost, 2020). Despite this, mental health interventions continue to be constructed from a deficit-focused lens.

National Institute for Health and Care Excellence, 2013 (NICE) guidance (National Institute for Health and Care Excellence, 2013) recommends that mental health conditions experienced by autistic people, such as anxiety problems, are managed using specifically adapted forms of evidence-based interventions. To date, adaptations have largely been based on theoretical models derived from non-autistic experiences in an attempt to make therapeutic interventions work for people with *Autism Spectrum Disorder* (as conceptualised by the pathologised view of autism). Indeed, the NICE guidelines reflect the current evidence, which has been created without meaningful involvement of autistic perspectives. NICE guidelines inform clinical practice, thus autistic people are being provided mental health interventions constructed from an exclusively non-autistic conceptualisation of autism. Adapted interventions currently available tend to have similar ingredients and although have been demonstrated to have some efficacy, are often viewed negatively by the autistic community. Most notably, Cognitive Behavioural Therapy, the most evidenced intervention for autistic young people experiencing mental health difficulties (Perihan et al., 2020) is often viewed as a poor fit for autistic people by those in the community (Adkin, 2022), predominantly for its epistemologically neuro-normative stance, which can encourage autistic people to mask and does not consider environmental goodness of fit. An alternative view comes from a neurodiversity-affirmative perspective, and considers the context of being autistic in a non-autistic world. The use of co-design approaches, involving the autistic community and experts by experience, is critical to developing neurodiversity-affirmative interventions that meet the needs of autistic people.

## The Programme Being Adapted

Anxiety problems are the most commonly occurring mental health difficulty for autistic people (Vasa et al., 2020), they have an early onset (Solmi et al., 2022) and can have life-long implications and negative impact both for the child and their family. Despite this, there is a significant gap in service provision for autistic children with anxiety problems.

In our initial discussions with parents of autistic children, they informed us that they often felt left alone to help their child and wanted to know how best they could help. A brief, therapist-guided, parent-led Cognitive Behaviour Therapy (CBT) for child anxiety problems was identified by the neurodiverse research team as a potential programme for co-adaptation.

Brief, guided parent-led CBT has been shown to effectively reduce anxiety problems in pre-adolescent non-autistic children (Creswell et al., 2017). This parent-led CBT for child anxiety problems has recently been transposed to a brief online version via a co-production process (Hill et al., 2022). The results have suggested that the online approach achieves good outcomes and can further increase access to treatment (Hill et al., 2022). Consistent with other findings that parent-based interventions which have been transposed to an online format have retained their efficacy whilst overcoming barriers to access (Prinz et al., 2022; Spencer et al., 2020). Thus, we have used evidence-based parent-led CBT content and the learnings from the online adaptation co-production process to co-produce an adapted parent-led treatment for child anxiety problems in the context of autism.

## Authors’ Position

The authors conceptualise autism from a neurodiversity lens and reject the traditional deficit focused model of autism. The focus of amelioration of symptoms is placed on those attributable to anxiety. In addition, there is a particular focus on promoting environmental adaptations and de-stigmatised views of autism to support the psychological health of autistic children.

## Funder’s Statement

This project was funded by the National Institute for Health and Care Research (NIHR) under its Research for Patient Benefit (RfPB) Programme (Grant Reference Number NIHR203495). The views expressed are those of the author(s) and not necessarily those of the NIHR or the Department of Health and Social Care. All experts by experience were paid for their involvement in the work. Pre-grant expert by experience work was funded via an NIHR Research Design Service Patient and Public Involvement grant.

## The current study

This study combined multi-perspective qualitative data with processes and strategies adapted from experience-based co-design (EBCD) to adapt parent-led CBT content to meet the needs of autistic children and their families. EBCD involves seeking insights from key stakeholder groups, in this case: parents, children, autistic adults, academics, clinicians (Donetto et al., 2015). Using these insights, we have created the key content for an online, brief, parent-led, anxiety intervention for autistic children (aged 5-12).

An embedded qualitative study used Interpretive Phenomenological Analysis, and Template Analysis to draw out key themes in relation to both children and parents’ experiences and understanding of anxiety problems among autistic children. The details of the qualitative study are not reported here (see Cullingham et al., n.d.), instead we describe below how the qualitative data contributed to the co-adaptation process. At the centre of the co-adaptation process was an Expert Reference Group (ERG).

## The Expert Reference Group (ERG)

The ERG consisted of parents (autistic and non-autistic) of autistic children with anxiety problems, an autistic young person (<18 years old) with experience of anxiety problems, and non-autistic clinicians with experience supporting autistic children with mental health difficulties.

## The Research Team

The research team consisted of a non-autistic patient and public involvement expert who is a parent of an autistic young man with longstanding anxiety problems, an autistic researcher who has research expertise in autism, particularly critical autism studies, who is also a parent of an autistic adult with a learning disability, a non-autistic consultant clinical psychologist and expert in child anxiety interventions, a non-autistic expert in qualitative methods and co-production methodology, a non-autistic consultant child and adolescent psychiatrist with expertise in autism science and clinical practice, and the lead author of this paper who is a non-autistic neurodivergent (dyslexic) clinical psychologist with clinical expertise in supporting autistic children, and their parents, struggling with mental health difficulties.

## Intervention Development Process

### Online Support and Intervention for Child Anxiety Intervention (OSI) for Non-Autistic Children

The original version of Online Support and Intervention for child anxiety (OSI), used to support non-autistic children, is an 8-session interactive web-based CBT intervention (including one welcome module and one follow-up module). The content supports parents to help their children overcome problems with anxiety through simple interactive and multi-modal content (Hill et al., 2022). Parents have weekly phone/online calls with a therapist to individualise the content (see HIll et al., 2022, supplementary material Table 1 for session summaries).

### Intervention Development

The research team worked alongside the ERG to adapt the parent-led CBT content from OSI. The NIHR Guidance on co-producing research (Hickey et al., 2018) as well as the Medical Research Council’s guidance (Skivington et al., 2021) on developing complex interventions was considered in developing the study. Using an iterative and reciprocal feedback process, we used insights from four sources: 1. academic literature; 2. qualitative data obtained from the embedded qualitative study; 3. the ERG; and 4. the research team; to create and check the acceptability of a neuro-affirmative, parent-led, anxiety intervention for autistic children aged 5-12.

An iterative feedback cycle between the research team and ERG was established in which insights were regularly shared, allowing both groups to consider and review the opinions of the other. Minutes were taken at every ERG meeting and key points summarised; the ERG members then checked these for accuracy. The minutes and summaries were shared with the research team after a 48-hour review period. Using deliberative practice, as is central to EBCD, the researchers translated the experiential knowledge gained from the professionals and experts by experience in the ERG into relevant adaptations to the intervention. The identified adaptations were then reviewed by the ERG for fact checking and consideration. This procedure was repeated throughout the adaptation process.

All information was stored on key insights documents which captured the learnings from all sources, and highlighted discrepancies between different sources.

The developing intervention content was written by the lead researcher, using the key insights documents, prioritising insights from the ERG and qualitative data. Written content was reviewed by the ERG. Any component of the adapted intervention that the ERG felt had the potential to do harm, was explored and discussed in detail. Following detailed discussion and problem-solving, if the ERG still expressed concerns this was recorded as not to be included within the adapted intervention. For further content on elements that the ERG identified as potentially harmful see themes from the ERG below.

If there was a disagreement between stakeholders the ultimate decision-making responsibility fell upon the lead researcher, considering insights and expertise from the research group and the ERG, prioritising insights from those with lived experience. However, this process was not needed as the ERG and the research team were able to agree on all content and no significant disagreements occurred.

The intervention development took place in three phases: study set-up; initial evidence gathering; evidence checking and content writing. See Figure 1 for a visual representation of intervention development.

**Figure 1.**
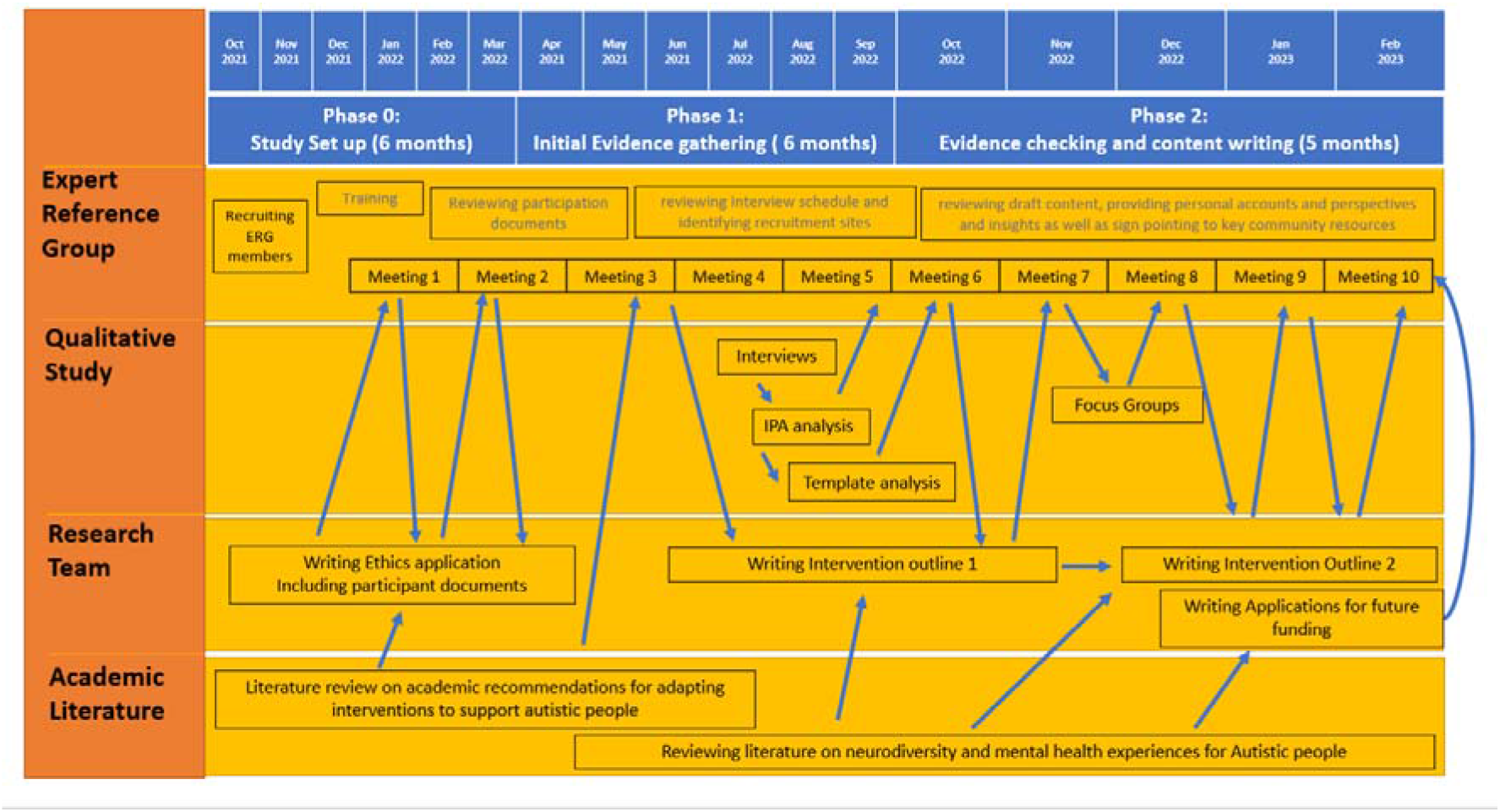
Visual representation of intervention development

### Phase 0: Study Set up

To establish the ERG, parent carer forums and autism charities sent out invitations for expressions of interest. The lead researcher and interested parties then had a conversation with potential members to discuss the role and answer any questions. Prior to the first meeting, ERG members were provided training and support.

The research ethics application for the embedded qualitative study and study documentation was created during this phase with input from the ERG.

Academic literature on anxiety problems experienced by autistic people (Vasa et al., 2020), effective interventions for autistic children with anxiety problems (Sharma et al., 2021; Spain & Happé, 2020), and the NICE guidelines (National Institute for Health and Care Excellence, 2013), were reviewed and synthesised by the research team.

### Phase 1: Initial Evidence Gathering

This phase consisted of gathering information and critically reviewing the information needed to adapt the intervention.

Findings from the academic literature were presented and discussed with the ERG. Additionally, the ERG members were asked to share their thoughts on what was needed to support autistic children with anxiety problems and their families. The opinions and reflections of the ERG were then taken back to the research team and integrated into the intervention.

During this phase, the embedded qualitative study was undertaken. Qualitative data were obtained via semi-structured interviews with 10 parents of autistic children with anxiety problems (children age 8-12 years, median age 10) and 9 autistic children with anxiety problems (children age 8-12 years, median age 10). Four parents participated in the focus groups, which were used to gain further insights into the acceptability of the draft content. The interviews were analysed using Interpretative Phenomenological Analysis (IPA) and Template Analysis. The themes from the qualitative analysis, input from academic literature (including NICE) and the ERG were combined on the key insights documents.

### Phase 2: Evidence Checking and Content Writing

The content of the intervention was developed in this phase. Using the key insights documents created during the evidence-gathering phase, a basic outline of the intervention and case examples were written and then critically reviewed with the ERG. Literature from Critical Autism studies (Heselton, 2021; Milton, 2018)) and from the neurodiversity movement (Dyck & Russell, 2020; Leadbitter et al., 2021) were explored and integrated into the intervention.

Once a clear intervention outline was established, this was taken and shared with focus groups consisting of parents of autistic children with anxiety problems to gain further opinions and insights into the acceptability of the intervention. These findings were shared with the ERG and research team and a final outline of the adapted intervention was established.

Funding plans and the future trial design was identified during this phase.

### ERG Meetings

The ERG met (remotely) monthly throughout the project to review all components of the adaptation and research process. ERG meetings and research team meetings were held separately. ERG meetings were co-chaired by the lead researcher and the co-investigator who brought lived experience of parenting an autistic child with anxiety problems to minimise inherent power imbalances and promote an atmosphere of open critique.

#### Training sessions

Two training sessions were held at the start of the study to explore required support and outline expectations. To promote a safe and familiar space and in acknowledgement of the typical power structures present in health care and research, separate training sessions were held for the clinicians and the non-clinical ERG members (those with lived experience). The ERG co-chairs thought it would be easier to openly discuss the power dynamics, perceived challenges, fears and worries about working together, and explore biases in self-contained groups. Members of the research team joined for part of both training sessions to introduce themselves and answer questions about the research process.

#### Co-constructed ERG structure

The ERG meeting structure was created by the ERG instead of being pre-conceived by the research team. This was established within the training sessions and initial meetings. All sessions had an informal agenda (sent out beforehand), regular breaks and check-ins. The meetings would open and close with reflections, with content shared via PowerPoint, with ample time for discussion and critique.

#### Facilitation of equitable communication

The ERG co-chairs met regularly throughout the project to reflect on their own chairing and how they were contributing to or dismantling power imbalances. The co-chairs decided to take on different roles within the ERG (for example the co-chair with clinical and academic experience would present the content and the co-chair with lived experience would lead the discussion to facilitate open and honest critique). Both co-chairs provided honest and unboundaried reflections during the reflection sessions to further dismantle power imbalances, sharing their worries/hopes about chairing such a group, as well as on-going challenges and successes of the project. The aim was to place equal expectations around honesty and sharing for all members of the ERG. Within the ERG efforts were made to create balanced communication, giving weight to all perspectives. This included providing options around communication with all communication styles being given the same importance whether verbal or written in the chat function, the option to have cameras on or off, and the use of online meeting features such as hands-up. At all meetings, an invitation to be critical was reiterated, and differences of opinion were openly discussed and celebrated.

### Objectives of ERG Meetings

#### Meeting one

The ERG decided on a structure for meetings. The plan included reflections during the first few minutes of the sessions and a shared decision about what digital platform to use. Group rules were decided together and shared (see supplementary material for group rules).

#### Meeting two

An overview of the parent-led CBT intervention and questions answered by one of the programme’s originators. Further review of documentation and ethics was completed.

#### Meeting three and four

The ERG critically reviewed a summary of empirical literature prepared by the research team on factors to consider when adapting neurotypical interventions to meet the needs of autistic individuals (Cooper et al., 2018; Walters et al,. 2016). See Table 1 for further information. Components were highlighted as being advantageous, neutral or deleterious.

**Table 1.**
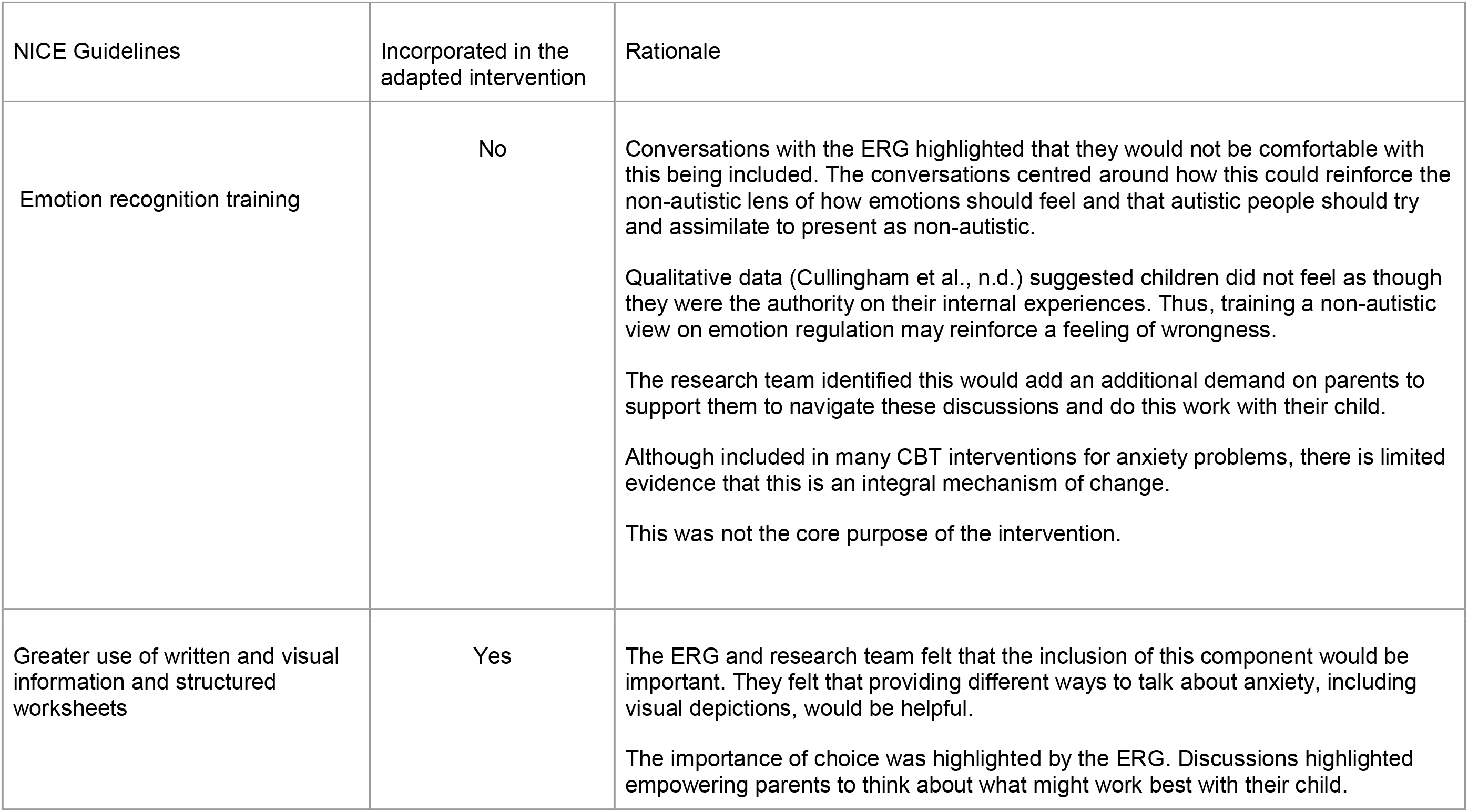
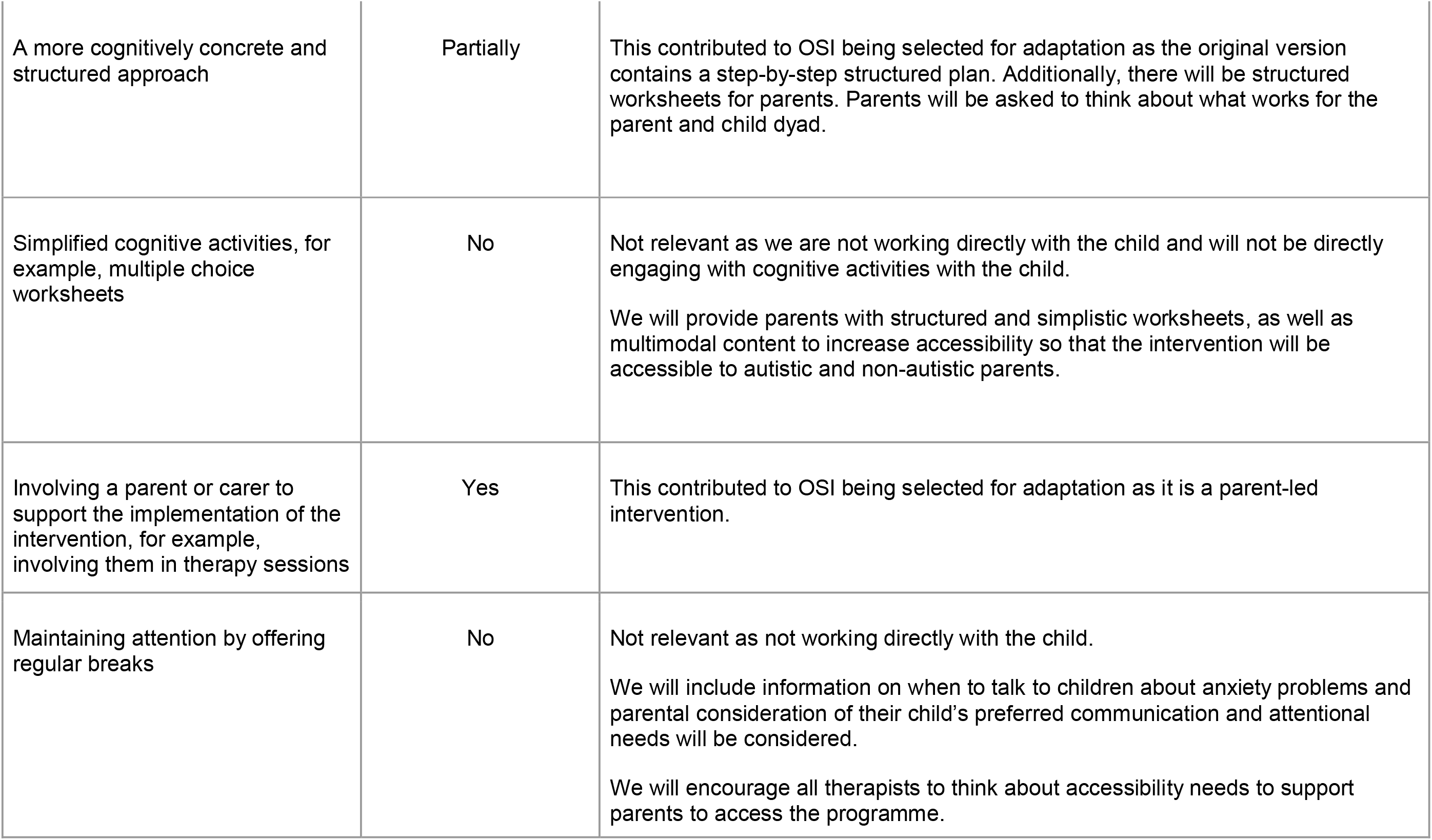
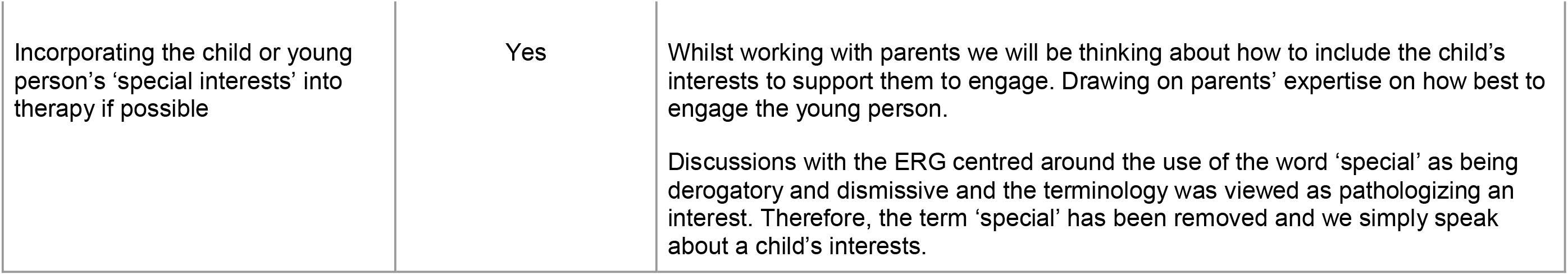
Elements from NICE guidance for amending interventions

#### Meeting five, six, seven and eight

The ERG was presented with data from the qualitative analysis for checking and sense making. The ERG also reviewed an iterative blueprint of the adapted intervention at each of these meetings.

#### Meetings nine and ten

The ERG continued to review the intervention draft content. Next steps were considered and ways to disseminate the findings as well as future funding applications were considered.

## Results

### Key Themes Extracted from the ERG Meetings

#### Autism-specific understanding and information

All ERG members felt strongly that without well-integrated neuro-affirmative psychoeducation adapted for the specific needs of autistic children, the intervention would likely not be optimally effective or acceptable for parents. There was a large focus on the importance of providing psychoeducation on how common autistic needs interface with anxiety problems. Throughout the iterative process, key areas of psychoeducation were identified, discussed and finalised. This included information on sensory needs and how they relate and do not relate to anxiety problems, interoception differences, autistic shutdown and overwhelm, the up-and-down nature of anxiety for autistic children, and understanding daily underlying stressors.

#### The ability for the programme to be personalised and tailored to individual needs

The ERG highlighted the importance of thinking about the individual child and the need for personalisation throughout the programme rather than a ‘one size fits all’ approach, and how this could be achieved within a brief programme. Two children may present similarly however one child’s presentation may reflect their anxiety whilst for the other it may reflect a sensory need, or a combination of both. ERG members reflected together on how hard it can be for the child, or parent, to understand the underlying elements and how best to support the child. The ERG identified that without bringing the content back to the individual child and their needs it would be easy to misinterpret elements of the child’s needs and resources. Peer support was highly valued; however, not at the expense of personalisation. Flexible delivery methods were discussed such as psychoeducation groups with individual sessions for the intervention implementation. However, consideration of implementation ease, access and personalisation led to the decision of mainlining the OIS delivery method of an interactive on-line programme with telephone support.

#### Validating and understanding the whole family’s experience

The ERG emphasised the importance of acknowledgement that the whole family, including siblings, are involved and impacted by the anxiety problem. Ideas around how to support families with this and provide information on self-care were discussed, however many ERG members stated self-care recommendations felt-patronising, and often didn’t feel possible. The conversation with the ERG highlighted that importance should be placed on helping parents support their child, whilst understanding and validating the challenges. Support to set realistic goals, giving parents a protected place to think about their child’s needs, discussing and supporting parents to think about whether the programme is right for them right now, and acknowledging the challenges and the risks, were viewed as helpful.

#### Pressure to conform is an additional stressor for autistic children and their families

The pressure of societal expectations and the push to do things in a neuro-conformative non-autistic way were discussed. ERG members expressed perceived and explicit pressures from school, internal expectations, and the wider family to have the child conform to expectations. Providing space in the intervention to help parents build their confidence in allowing their child to do things in their own way was highlighted. Providing space for parents to think and reflect on questions such as “Does my child need to do this?”. “Will it help them in later life?”, “Is it the right thing to focus on to help them overcome their anxiety problems?”.

#### Potential elements that could do harm

The ERG expressed that without thinking about the autistic needs of a young person, and considering environmental goodness of fit, CBT could be used in a harmful way, as they may be a risk it could be used to encourage children to mask. This was managed by including psychoeducation that moved away from pathologising language, and highlighting the challenges of being autistic in a non-autistic world. The ERG members reflected on the importance of a neurodiversity lens, ensuring that it was clearly communicated that the aim of the programme was not to change or challenge a child’s autistic elements, but to give parents the tools to support their children manage their anxiety problems. The ERG stressed that parents would need support to distinguish between their child’s autistic needs and their anxiety problems, to decide what areas to focus on, using the graded exposure ladder.

The use of strategies created from a non-autistic lens were considered to potentially do more harm than benefit. The ERG expressed concerns that these strategies could potentially reinforce narratives that the autistic way of thinking was wrong and children should try to think and feel more like their non-autistic peers. Thus we have not included specific relaxation techniques created from a non-autistic lens or emotional labelling/emotional literacy work.

### Recommendations from the literature

The academic literature makes several recommendations for key adaptations that can be used to adapt CBT interventions to meet the needs of autistic people. It appears that the majority of these recommendations aim to enable autistic people to access ‘neurotypical’ approaches to treatment, they have not been created by, or in consultation with, autistic people and there appears to be little direct evidence of their benefits. See Table 1 for a review of NICE guidelines (National Institute for Health and Care Excellence, 2013) for amending interventions, which ones were included, which were not, and the reasons for those decisions.

## Key Adaptations

### Programme overview

As shown in table 2, the adapted programme has 8 content sessions, an introduction session and check-in session, resulting in a 10-session online programme. Similar to the original OSI, parents work through an online module, which includes written content, video testimonials, animations and interactive worksheets. Following review of the module, parents have a coaching session with a therapist (trained in the programme) to problem solve, review and personalise the content. While longer than the original (8 session) intervention, the adapted intervention remains light touch with less than 5 hours of therapist contact time and is largely self-directed.

**Table 2.** Intervention summary

| Intervention summary |  |
| --- | --- |
| OSI -ASD | Summary |
| **Welcome** (session 0) | Introduction session to address parents’ potential concerns about participating. Information on the stance and aim of the programme as well as barriers to change. |
| **Session 1**<br>Get ready: Anxiety | Information is provided about the impacts of anxiety, typical anxiety cycles, and information on why autistic children are |
|  | more prone to anxiety problems. Anxiety problems are depicted in terms of anxious thoughts and feelings as well as recognising contributing environmental factors. Parents are introduced to the idea of underlying stressors draining a child's resource battery. |
| **Session 2**<br>Get ready: Underlying stressors and environmental changes | Information on underlying stressors and environmental changes are presented. Common sensory and environmental strategies are presented. The session explores the parallel process of thinking about adaptations whilst applying strategies to overcome anxiety problems. Parents are asked to think about the support their child may need and things that help charge their child's resource battery. |
| **Session 3**<br>Understanding what keeps your child's anxiety going | This session allows for constructing a 'My child's anxiety' formulation reflecting the contribution of thoughts, behaviours and environmental factors. |
| **Session 4**<br>Have a go thinking | This session helps parents think about how they will talk to their child about overcoming anxiety problems. Information on different types of communication strategies is presented as well as information on autistic shutdown and overwhelm. |
| **Session 5**<br>Facing fears | Information on how to implement a graded exposure ladder is presented alongside environmental adaptations to promote adaptive learning. |
| Review and check in session | No content is provided. Check in to review implementation of graded exposure ladder and review content as needed. |
| **Session 6**<br>Becoming independent | Supports parents to think about encouraging independence in a way that is appropriate for their child including information about their potentially unpredictable/ uneven developmental trajectory. |
| **Session 7**<br>Problem solving | Module focuses on problem-solving using CBT strategies. |
| **Session 8**<br>Keep it going | Promotes parents to plan for the future and to continue to make changes following the programme. |
| Follow up session | Phone call to review |

The adapted programme invites parents to think about the needs of their child within the context of their social (predominantly non-autistic) world and ways in which autistic styles of thinking and understanding can contribute to developing and maintaining anxiety problems. The way in which autism is defined and constructed and the language around autism has been carefully selected with PPI representatives to promote a non-pathologizing view of autism. The psychoeducation component of the programme has been extended and re-organised to meet the needs of autistic children. Throughout the intervention, importance is placed on thinking about ways to mitigate daily stressors and environmental adaptations that may be needed to help a child face their fears in a way that will promote adaptive learning. Information has been added on ideas for environmental adaptations, sensory strategies for emotional regulation, and goal setting around these. Parents are encouraged to work through the content and personalise it to the specific needs of their child.

The graded exposure ladder now has environmental adaptations placed alongside it. We have also added information on “charging your child’s resource battery” and asked parents to think about things that drain their child’s resource battery, to help parents understand how overwhelming a child’s experience can be. An additional check in session has been added to help parents’ problem solve and think about their child’s unique needs and the environmental goodness of fit. The independence session invites parents to think about their children’s unique developmental profile.

## Discussion

Co-production has been fundamental to this research having epistemological integrity and the resulting output holding a neurodiversity-affirmative stance. The involvement of people with lived experience in all stages of intervention development has allowed the integration of multiple and sometimes contrasting perspectives.

Throughout the process productive debates around constructs and held assumptions allowed for the open exploration of what was needed in the intervention, what was harmful and what could be done differently. We hope that co-producing this intervention has prevented it from inadvertently furthering internalised stigma. This process has allowed the team to move away from language and therapeutic techniques that can reinforce ableist beliefs that the way autistic children think and feel is wrong. We have thoughtfully selected the therapeutic ingredients that we believe will support autistic children to overcome anxiety problems, whilst celebrating their differences, strengths and acknowledging the challenges they face. There was a strong sense from participants that this approach ensured language used and the intervention itself were in-keeping with a neurodiversity-affirmative approach.

### Challenges and Learnings about Co-production

There were several challenges and limitations of this project and more work is needed to create clearer and more effective ways of co-producing mental health interventions with autistic people. As is well documented in the co-production literature, co-production is not without limitations or challenges. The study, particularly the set-up, was more time-consuming than if experts by experience had not been integrated into the design (Oliver et al., 2019).

The iterative nature of the project as well as integrating multiple perspectives took longer than considering only the academic literature and traditional academic networks. This can be overcome by careful planning and allowing for longer lead times to facilitate engagement. Working alongside experts by experience from the beginning is integral to successfully a co-production project, from both an epistemological integrity standpoint and a practical standpoint. Beyond their expert and intimate subject knowledge, experts by experience add invaluable knowledge about facilitating non-tokenistic engagement, supporting non-academics to feel able to contribute, and develop training materials for all members of the team (researchers and experts by experience alike).

Another challenge of co-production is incorporating many different experiences. researchers and experts by experience can bring and often do bring their own biases. We think this is a strength of the project as by having stakeholders with differing opinions we hope we have challenged many biases researchers and experts by experience may not have been aware of.

It is important to note that a limitation of this project was the ethnicity, socioeconomic status, sexuality/gender and cultural diversity within the ERG; Although a neurodiverse and experientially diverse group, the ERG was predominantly white British. People from communities that are an ethnic minority in the UK are under-represented in research, as are Autistic voices. Including representatives of intersectional experience, such as perspectives from multiple ethnic cultures would have enhanced this project. More work is needed to further promote the involvement of ethnically diverse co-production within the field of autism.

### Future Research

More research and development of co-production with neurodivergent populations is needed to support the development of more inclusive, sophisticated, and neuro-affirmative interventions. Within the academic community there is still hesitation at times about whether the benefits of co-production outweigh the costs. Although theoretically robust, the resulting co-produced interventions still need efficacy and effectiveness testing to fully understand the outcomes and potential benefits of co-production within mental health interventions.

This research has presented a systematic and fully experience-based co-design methodology within a sensitive area of child mental health care, making this study an important benchmark for methods going forward. Through this methodology we have created a novel intervention that integrates multiple theoretical backgrounds and is potentially acceptable to clinicians, autistic people and their family members. The co-constructed intervention adheres to a Cognitive Behavioural Therapy approach, whilst integrating ideas and concepts from the neurodiversity movement, thus establishing a mental health intervention that is built on empirical evidence but moves away from a pathologising view of autism.

##### Key points

###### What is known

Co-production is often recommended yet, to date there are limited methodological papers that explore how to successfully co-produce mental health interventions with autistic people.

###### What is New

This paper provides a methodological example of successful co-production within a sensitive area of child mental health promoting the creation of more neuro-affirmative mental health interventions

###### What is relevant

The resulting co-constructed intervention appears to adhere to Cognitive behavioural techniques, whilst integrating ideas and concepts from the neurodiversity movement. Thus, establishing a mental health intervention that is built on empirical evidence but moves away from a pathologizing view of autism

## Data Availability

Data is not available due to confidentiality

## Acknowledgements

In addition to the funders and co-authors we would like to acknowledge the contributions of the assistant psychologists and members of the ERG (non-authors) who made this project possible: Victoria Taylor, Bec Lee, Joanne Parry, and Ella Solomon. We would also like to acknowledge and remember the contributions of the late Dr Kishan Sharma.

## Funder’s Statement

This project was funded by the National Institute for Health and Care Research (NIHR) under its Research for Patient Benefit (RfPB) Programme (Grant Reference Number NIHR203495). The views expressed are those of the author(s) and not necessarily those of the NIHR or the Department of Health and Social Care.

All experts by experience were paid for their involvement in the work. Pre-grant expert by experience work was funded via an NIHR Research Design Service Patient and Public Involvement grant.

## Notes

### Competing Interest Statement

The authors have declared no competing interest.

### Author Declarations

Manchester Foundation Trust was the sponsor. The project was approved by the HRA (IRAS 307932), and Funded by via NIHR RFPB (NIHR203495)

## References

Adkin, D. G.-H. A. (2022, January 2). *Creating Autistic Suffering: Neuronormativity in Mental Health Treatment*. Emergent Divergence. https://emergentdivergence.com/2022/01/02/creating-autistic-suffering-neuronormativity-in-mental-health-treatment/

Botha, M., & Frost, D. M. (2020). Extending the Minority Stress Model to Understand Mental Health Problems Experienced by the Autistic Population. Society and Mental Health, 10(1), 20–34. https://doi.org/10.1177/2156869318804297

Creswell, C., Violato, M., Fairbanks, H., White, E., Parkinson, M., Abitabile, G., Leidi, A., & Cooper, P. J. (2017). Clinical outcomes and cost-effectiveness of brief guided parent-delivered cognitive behavioural therapy and solution-focused brief therapy for treatment of childhood anxiety disorders: a randomised controlled trial. The Lancet. Psychiatry, 4(7), 529–539. https://doi.org/10.1016/s2215-0366(17)30149-9

Cullingham, T., Larkin, M., Milton, D., Rennard, U., Taylor, V., Solomon, E., Green, J., & Creswell, C. (n.d.). Understanding experiences of anxiety for autistic children and their parents. https://www.medrxiv.org/

Donetto, S., Pierri, P., Tsianakas, V., & Robert, G. (2015). Experience-based Co-design and Healthcare Improvement: Realizing Participatory Design in the Public Sector. The Design Journal, 18(2), 227–248. https://doi.org/10.2752/175630615X14212498964312

Dyck, E., & Russell, G. (2020). Challenging Psychiatric Classification: Healthy Autistic Diversity and the Neurodiversity Movement. In S. J. Taylor & A. Brumby (Eds.), Healthy Minds in the Twentieth Century: In and Beyond the Asylum (pp. 167–187). Springer International Publishing. https://doi.org/10.1007/978-3-030-27275-3_8

Heselton, G. A. (2021). Childhood adversity, resilience, and autism: a critical review of the literature. Disability & Society, 1–20. https://doi.org/10.1080/09687599.2021.1983416

Hickey, G., Brearley, S., Coldham, T., Denegri, S., & Green, G. (2018). Guidance on co-producing a research project. Southampton: INVOLVE.

Hill, C., Reardon, T., Taylor, L., & Creswell, C. (2022). Online Support and Intervention for Child Anxiety (OSI): Development and Usability Testing. JMIR Formative Research, 6(4), e29846. https://doi.org/10.2196/29846

Leadbitter, K., Buckle, K. L., Ellis, C., & Dekker, M. (2021). Autistic Self-Advocacy and the Neurodiversity Movement: Implications for Autism Early Intervention Research and Practice. Frontiers in Psychology, 12, 635690. https://doi.org/10.3389/fpsyg.2021.635690

Milton, D. (2018). Autistic Development, Trauma and Personhood: Beyond the Frame of the Neoliberal Individual. In K. Runswick-Cole, T. Curran, & K. Liddiard (Eds.), The Palgrave Handbook of Disabled Children’s Childhood Studies (pp. 461–476). Palgrave Macmillan UK. https://doi.org/10.1057/978-1-137-54446-9_29

National Institute for Health and Care Excellence. (2013). Autism spectrum disorder in under 19s: support and management (Clinical guideline [CG170]). https://www.nice.org.uk/guidance/cg170/chapter/Recommendations#interventions-for-coexisting-problems

Oliver, K., Kothari, A., & Mays, N. (2019). The dark side of coproduction: do the costs outweigh the benefits for health research? Health Research Policy and Systems / BioMed Central, 17(1), 33. https://doi.org/10.1186/s12961-019-0432-3

Perihan, C., Burke, M., Bowman-Perrott, L., Bicer, A., Gallup, J., Thompson, J., & Sallese, M. (2020). Effects of Cognitive Behavioral Therapy for Reducing Anxiety in Children with High Functioning ASD: A Systematic Review and Meta-Analysis. Journal of Autism and Developmental Disorders, 50(6), 1958–1972. https://doi.org/10.1007/s10803-019-03949-7

Prinz, R. J., Metzler, C. W., Sanders, M. R., Rusby, J. C., & Cai, C. (2022). Online-delivered parenting intervention for young children with disruptive behavior problems: a noninferiority trial focused on child and parent outcomes. Journal of Child Psychology and Psychiatry, and Allied Disciplines, 63(2), 199–209. https://doi.org/10.1111/jcpp.13426

Sharma, S., Hucker, A., Matthews, T., Grohmann, D., & Laws, K. R. (2021). Cognitive behavioural therapy for anxiety in children and young people on the autism spectrum: a systematic review and meta-analysis. BMC Psychology, 9(1), 151. https://doi.org/10.1186/s40359-021-00658-8

Skivington, K., Matthews, L., Simpson, S. A., Craig, P., Baird, J., Blazeby, J. M., Boyd, K. A., Craig, N., French, D. P., McIntosh, E., Petticrew, M., Rycroft-Malone, J., White, M., & Moore, L. (2021). A new framework for developing and evaluating complex interventions: update of Medical Research Council guidance. BMJ, 374, n2061. https://doi.org/10.1136/bmj.n2061

Solmi, M., Radua, J., Olivola, M., Croce, E., Soardo, L., Salazar de Pablo, G., Il Shin, J., Kirkbride, J. B., Jones, P., Kim, J. H., Kim, J. Y., Carvalho, A. F., Seeman, M. V., Correll, C. U., & Fusar-Poli, P. (2022). Age at onset of mental disorders worldwide: large-scale meta-analysis of 192 epidemiological studies. Molecular Psychiatry, 27(1), 281–295. https://doi.org/10.1038/s41380-021-01161-7

Spain, D., & Happé, F. (2020). How to Optimise Cognitive Behaviour Therapy (CBT) for People with Autism Spectrum Disorders (ASD): A Delphi Study. Journal of Rational-Emotive and Cognitive-Behavior Therapy: RET, 38(2), 184–208. https://doi.org/10.1007/s10942-019-00335-1

Spencer, C. M., Topham, G. L., & King, E. L. (2020). Do online parenting programs create change?: A meta-analysis. Journal of Family Psychology: JFP: Journal of the Division of Family Psychology of the American Psychological Association, 34(3), 364–374. https://doi.org/10.1037/fam0000605

Vasa, R. A., Keefer, A., McDonald, R. G., Hunsche, M. C., & Kerns, C. M. (2020). A Scoping Review of Anxiety in Young Children with Autism Spectrum Disorder. Autism Research: Official Journal of the International Society for Autism Research, 13(12), 2038–2057. https://doi.org/10.1002/aur.2395

